# Enteral Vs Oral Nutrition in non-interrupted compliance with chemoradiotherapy in subjects with esophageal cancer

**DOI:** 10.1101/2023.09.15.23295559

**Authors:** Ayder Chacón Corena, Hernando Chacón Chacón, Rusvelt Vargas Moranth, Grey Sierra Maury

## Abstract

**Background:** Esophageal cancer (EC) generates malnutrition, added to the adverse effects of chemoradiotherapy. It is important to avoid intratreatment interruptions to improve survival. The objective is to evaluate the effect of Enteral Nutrition (EN) Vs Oral Nutrition (ON) in compliance without interruptions of chemoradiotherapy in adults with esophageal cancer, in two centers of La Plata Argentina, during 2014-2019.

**Methods:** Retrospective cohort study. 76 subjects participated, 34 with EN and 46 with ON. Independent variable: type of nutrition, dependent variable: compliance with chemoradiotherapy; potentially confusing variables: sex, age, comorbidities, risk factors, and cancer stages. The percentages of qualitative variables were compared through Chi2 and the T test for the quantitative ones. The RR (95% CI) was calculated by comparing the interruptions in both groups. A crude and adjusted RR analysis was performed, stratifying by sex and clinical variables.

**Results:** 34 subjects had EN and 42 ON; mean age: 65.9 (SD +/-: 10.5) and 68.4 (SD +/-: 9.9) respectively. In both groups, most participants with interruptions were men, in advanced stages, with radical treatment and a diagnosis of carcinoma. In the EN group, 44.12% had interruptions, compared to 14.3% of ON (p <0.05). The group with EN interrupted a mean of 9.9 days (SD +/-: 5.9) and ON: 8.3 (SD +/-: 9.2) (p> 0.05).

**Conclusion:** The group with EN presented more interruptions, therefore it is not recommended for use prophylactically, only if necessary.

## Introduction

Esophageal cancer (EC) is one of the diseases that has a negative impact on human nutrition, leading to malnutrition in patients, in turn increasing the morbidity and mortality of individuals^1^. Malnutrition due to deficit, which occurs when there is a weight loss greater than 10% and / or with a serum albumin concentration less than 30 g / L^2^ is caused by several factors such as dysphagia, odynophagia, early satiety, cough caused by the presence of the tumor, anxiety, depression and loss of appetite^3^. It is advisable to carry out a nutritional screening prior to and during radiotherapy (RT) treatment to identify and correct the nutritional deficit, as well as maintaining and preserving weight, since it has been shown that malnutrition is present in 79% of cases before starting treatment^3,4^ which hinders the positive response to different cancer treatments and affects the quality of life in patients^4^.

Neoadjuvant chemoradiation treatment for approximately 5.5 weeks followed by surgery 7 weeks later is the choice for patients with locally advanced or node-positive esophageal squamous cell carcinoma^5,6^. The ChemoRadiotherapy for Oesophageal Cancer followed by Surgery Study (CROSS) confirmed the survival benefit for preoperative chemoradiation with Paclitaxel and Carboplatin in patients with resectable esophageal or gastroesophageal junction cancer. The preferred preoperative regimen is Paclitaxel (50 mg / m2) and intravenous carboplatin (on days 1, 8, 15, 22 and 29) concurrent with external radiation therapy (EBRT) with a total dose of 41.4 Gy in 23 fractions of 1.8 Gy, 5 fractions per week^6,7^. Individuals who are not surgical candidates or who refuse surgery can be treated with definitive chemoradiation^5^. Preoperative RT doses range from 41.4–50.4 Gy (1.8–2.0 Gy / d), the postoperative dose range is 45–50.4 Gy (1.8–2.0 Gy / d) and the definitive RT dose is 50–50.4 Gy (1.8–2.0 Gy d), higher doses (60–66 Gy) may be appropriate for cervical esophageal tumors when surgery is not planned ^7^.

Chemoradiotherapy can contribute to the improvement of dysphagia by up to 50%, taking weeks to take effect. Among the adverse effects produced by the treatment, weight loss, catabolism, stenosis, mucositis, odynophagia and radioinduced pneumonia may occur. Therefore, the important to start with an initial nutritional diagnosis and adequate support from the moment a cancer diagnosis is made to improve treatment outcomes and patient quality of life^1,2,4.^

Weight loss during chemoradiotherapy is associated with the lack of nutritional advice, stages III and IV according to TNM, total energy intake <1441.3 kcal / day, depression, esophagitis and loss of appetite^6^. When patients cannot consume 50-75% of their requirements through diet for more than 5 consecutive days or are malnourished, oral nutritional supplements should be administered^1^. If the oral food intake is insufficient, enteral nutritional support (EN) with standard formulas is recommended according to the specific needs of the disease, the purpose is to increase the intake of macro and micronutrients directly in the gastrointestinal (GI) tract through of probes without passing through the oral cavity^8^. Preoperatively, artificial nutrition is recommended for severely malnourished (weight loss> 20%) and moderately malnourished (10-19% weight loss) who will undergo surgery^1^. The nasogastric tube (NGT) / nasojejunal (NJ) is prescribed for a duration of less than 30 days in patients and for those who need a duration greater than 30 days, percutaneous endoscopic gastrostomy (PEG) / percutaneous endoscopic jejunostomy (PEJ) is indicated, except for those that require Preoperative RT^3^.

A retrospective cohort study in patients with EC, under multimodal therapy in conjunction with EN, concluded that there is a potential increase in the risk of mortality with this type of nutrition^9^. Regarding the quality of life of patients undergoing chemoradiotherapy in relation to EN, Yu et al.,^10^ compared NGT, esophageal stent and ostomies with feeding tubes, concluding that there is slightly worse quality of life in patients with esophageal squamous cell carcinoma using NGT, compared with the percutaneous route during chemoradiotherapy. The group with NGT presented greater pain due to its insertion worsening odynophagia.

When normal oral or modified nutrition, with supplementation, cannot ensure sufficient nutritional and energy intake or when it is predicted that the energy intake will be <60% of the expected energy use for more than 10 days, for a patient with a functioning digestive system, EN should be started. When a tracheoesophageal fistula occurs and adverse effects of chemotherapy (CT) and RT such as vomiting and diarrhea make oral feeding impossible^11^. EN in patients receiving preoperative CT or chemoradiotherapy, benefits them in maintaining weight, reducing toxicity and preventing treatment interruption^12^, which is key, since it has been shown that treatment interruptions in people with cancer decrease overall survival^13^.

The guidelines of the European Society for Clinical Nutrition and Metabolism (ESPEN) on nutrition in cancer patients recommend an energy requirement of 25-30 kcal / kg / day, protein intake of 1-1.5 g / kg / day, supplemented with electrolytes and the use of a multivitamin-multimineral supplement in doses equivalent to the recommended daily amount, making it a useful and safe measure^14^. Oral nutritional supplements (ONS) reduce side effects and improve clinical outcomes in cancer treatment. One study showed that ONS can reduce weight loss and improve the nutritional status of esophageal cancer patients during radiotherapy^15^. According to a systematic review in cancer patients who received CT or chemoradiation, oral glutamine significantly reduced the incidence of mucositis and the maximum degree of mucositis^12^.

The objective of this study is to evaluate the effect of EN Vs ON in the uninterrupted compliance of chemoradiotherapy in patients aged 46 to 93 years with esophageal cancer, in two RT centers in La Plata Argentina, from 2014-2019.

## Materials and Methods

A retrospective cohort design was applied. The study was conducted in two RT clinics in La Plata Argentina, with subjects who were seen from January 2014 to December 2019. The study began on November 1, 2019 and ended on June 30, 2020. 101 subjects participated with esophageal cancer at any esophageal level, between 46 and 93 years old, who underwent RT, with staging (E) by TNM I-IV, under treatment with chemoradiotherapy. 76 subjects were included and 25 subjects were excluded, of which 16 abandoned treatment due to severe complications and 9 who died during chemoradiotherapy. Interruption was considered to be the loss of one or more consecutive RT sessions in the week. It was found that 34 subjects had received EN and 42 ON. The variables studied were: type of nutrition (independent variable) and compliance with chemoradiotherapy treatment (dependent variable). The potentially confusing variables were: sex, age, comorbidities, risk factors, and cancer stages.

The data collection was carried out by searching the clinical records of each subject, with previous request to obtain permission from the RT institutions and later evaluation of a bioethics committee to obtain the endorsement of the use of confidential information, under the anonymity of the participants. For data collection, an Excel file was made, where the information was classified for statistical analysis. The percentages of clinical and sociodemographic variables were compared through Chi2 for the qualitative ones and the T test for the quantitative ones. Likewise, the RR (95% CI) was calculated by comparing the interruptions in both groups, and a crude and adjusted RR analysis was performed by stratifying sex and clinical variables (the latter dichotomized as can be seen in Table 2).

This work is framed within the Declaration of Helsinki of the World Medical Association and the Declaration of Geneva, where the fundamental principles of confidentiality, beneficence, non-maleficence, justice and respect for the autonomy of the participants are highlighted^16,17^.

## Results

In the group that underwent the EN treatment (n = 34), 4 had PEG, 27 NGT and 3 NJ. The most frequent type of EN was NGT with 79.4%. The mean age was 65.9 and 68.4 in the EN and ON groups, respectively. Most of the participants were men: 70.6% (EN) and 66.7% (ON). The predominant clinical stage was II: 47.1% (EN) and 40.5% (ON); the majority had radical treatment: 85.3% (EN) and 78.6% (ON), the most frequent diagnosis was carcinoma: 67.6% (EN) and 71.4% (ON) and the esophageal level with the highest percentage was the lower thoracic: 47.1% (EN) and 64.3% (ON). In none of the cases were the differences significant (p> 0.05), so the groups were comparable according to these variables (Table 1).

**Table 1.** Sociodemographic and clinical characteristics of the participants.

|  |  | %EN (n=34) | %ON (n=42) | p |
| --- | --- | --- | --- | --- |
| **Age (Mean; SD+/-)** |  | 65,9 (10,5) | 68,4 (9,9) | 0,293* |
| **Male sex** |  | 70,6 | 66,7 | 0,906** |
| **Stage** | **I** | 2,9 | 16,7 | 0,349** |
|  | **II** | 47,1 | 40,5 |  |
|  | **III** | 41,2 | 31,0 |  |
|  | **IV** | 21,4 | 35,7 |  |
| **Intention of treatment** | **Adyuvant** | 2,9 | 9,5 | 0,715** |
|  | **Neoadyuvant** | 8,8 | 9,5 |  |
|  | **Palliative** | 2,9 | 2,4 |  |
|  | **Radical** | 85,3 | 78,6 |  |
| **Histopathologic Diagnosis** | **Carcinoma** | 67,6 | 71,4 | 0,937** |
|  | **Adenocarcinoma** | 29,4 | 26,2 |  |
|  | **Other diagnoses** | 2,9 | 2,4 |  |
| **Esophageal level** | **Lower thoracic** | 47,1 | 64,3 | 0,786** |
|  | **Middle thoracic** | 29,4 | 16,7 |  |
|  | **Middle/lower thoracic** | 11,8 | 4,8 |  |
|  | **Upper / middle thoracic** | 5,9 | 4,8 |  |
|  | **Cervical** | 2,9 | 0,0 |  |
|  | **Upper thoracic** | 2,9 | 9,5 |  |
| **Type of EN** | **NGT** | 79,4 | - | - |
|  | **PEG** | 11,8 | - |  |

|  | %EN (n=34) | %ON (n=42) | p |
| --- | --- | --- | --- |
| NJT | 8,8 | - |  |

NGT: Nasogastric tube; NJT: Nasojejunal tube; PEG: Percutaneous endoscopic gastrostomy; \*T test; \*\* Chi2.

**Table 2.**
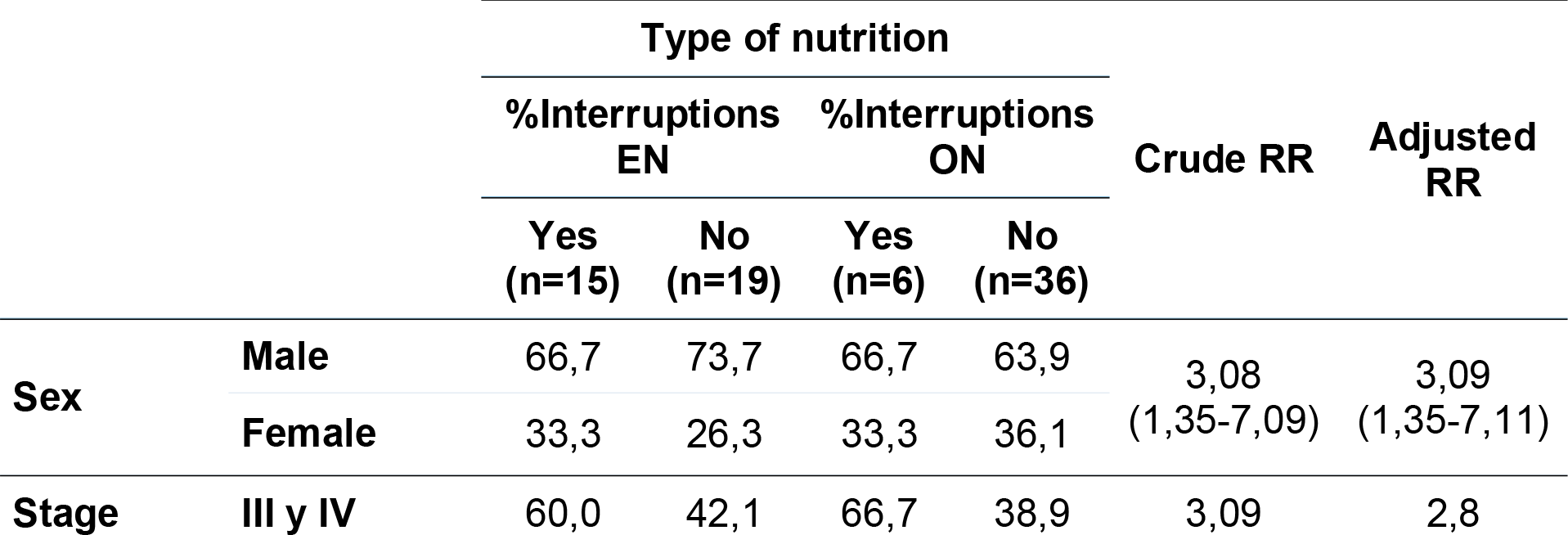
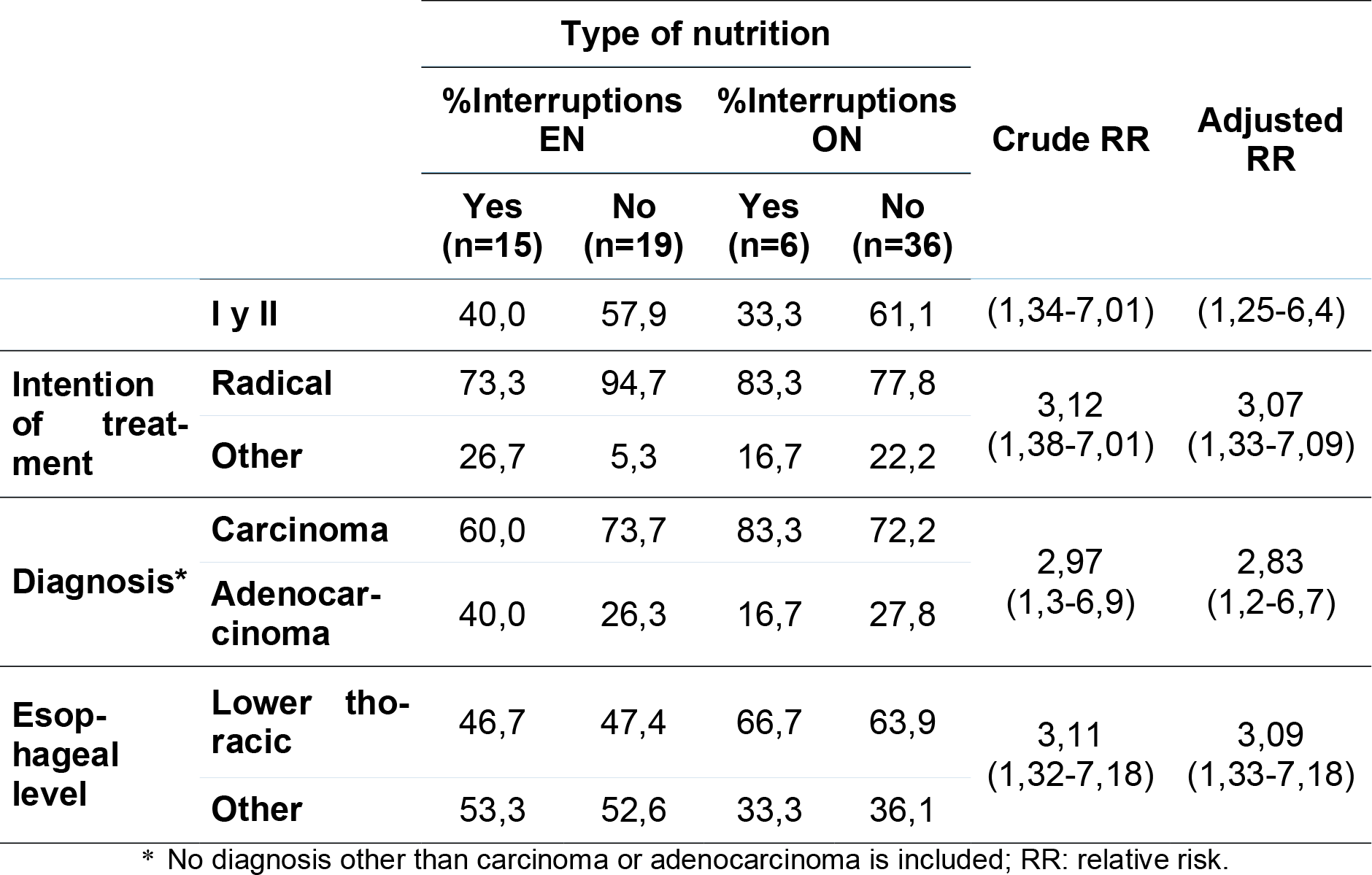
Percentage of subjects who make interruptions according to type of nutrition, clinical variables and sex.

On the other hand, the EN group had more than 2 times the risk of interrupting treatment than the oral group, significantly (p <0.05), since 44.12% of those belonging to the first group presented interruptions, compared to 14,3% of the second (Figure 1). The EN group interrupted an average of 9.9 days (SD +/-: 5.9) and ON: 8.3 (SD +/-: 9.2), these differences being not significant (p = 0.361).

**Figure 1.**
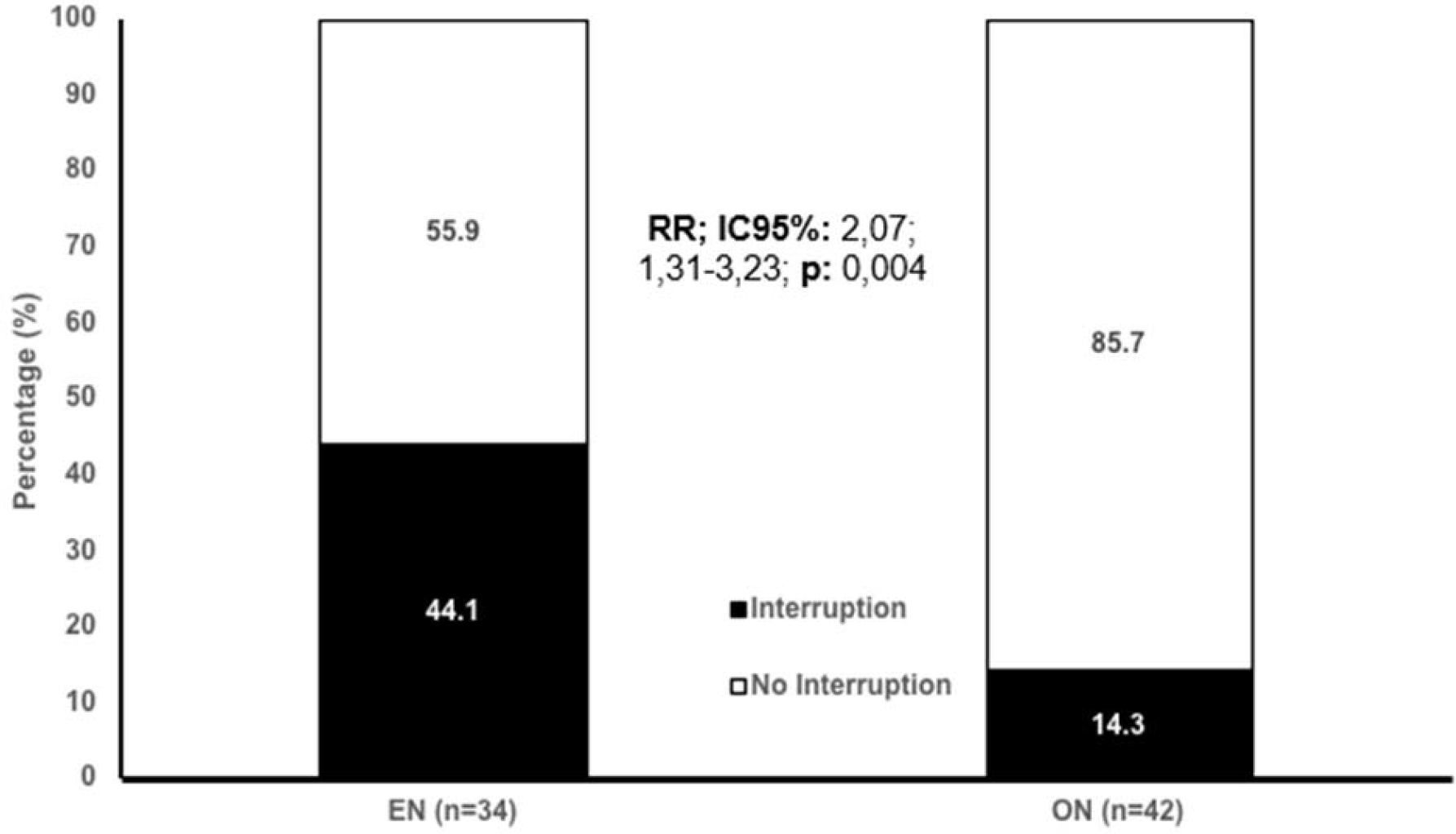
Percentage of subjects who interrupted treatment according to type of nutrition.

Within the subjects who received EN, 50% in which the type was PEG presented interruptions, as well as 44.4% of the NGT and 33.3% of the NJT, but these differences were not significant (p>0.05) (Figure 2).

**Figure 2.**
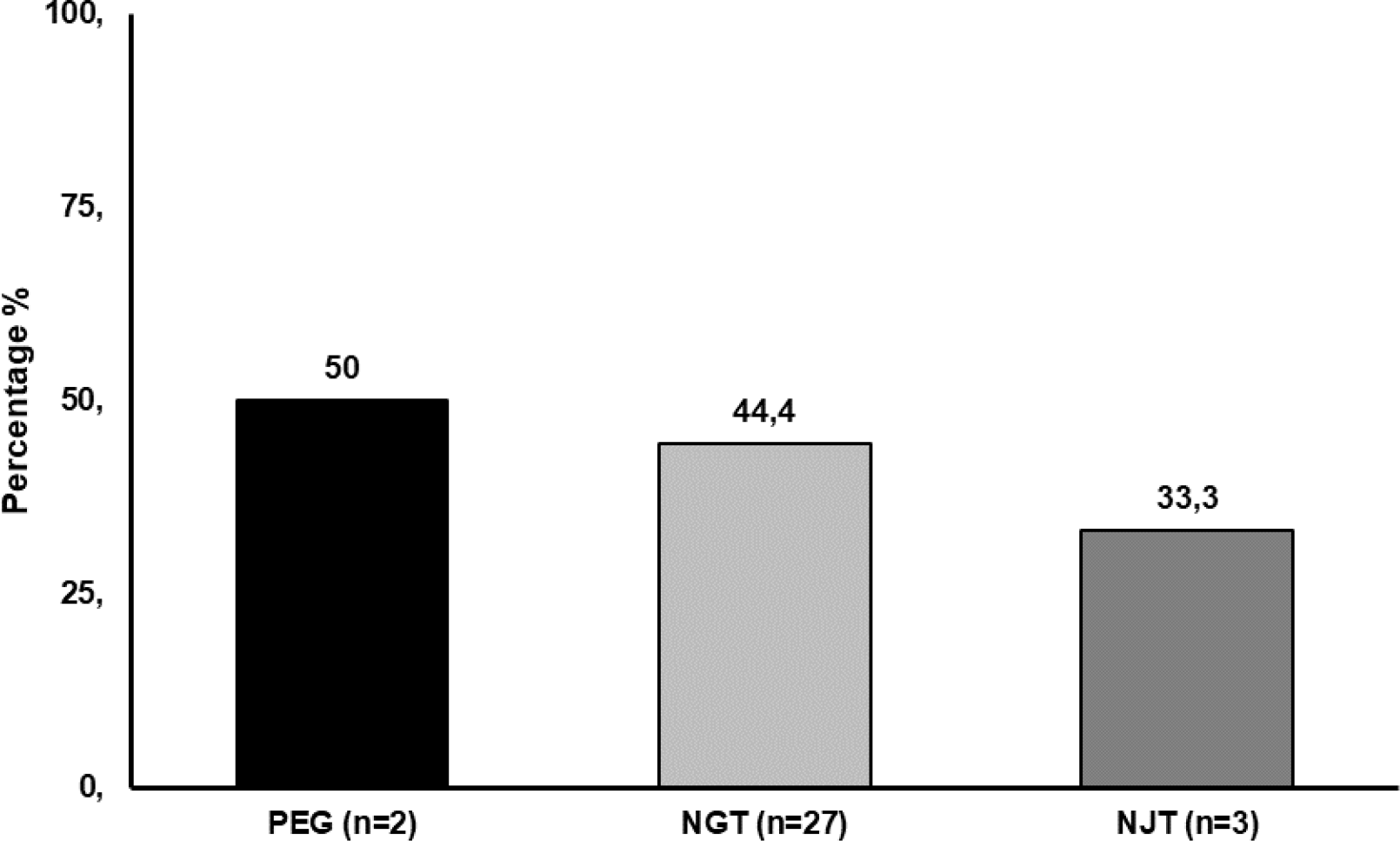
Percentage of subjects who had interruptions according to type of EN.

Among the participants who had interruptions, both in the EN and ON groups, the majority were men, in advanced stages, with radical treatment and a diagnosis of carcinoma; in reference to the esophageal level, in the ON group most of the interruptions were located in the lower thoracic and in the EN group they were located on a different level. In this stratified analysis the differences were significant (p <0.05), and the differences between the crude RR (3.09) and the adjusted one (2.8) suggest that only the stage could become a confounding variable, since the difference was very close to 10%, so in general terms, it could be said that the effect of the interruptions was mainly related to the type of nutrition, independently from the other variables (Table 2).

## Discussion

During RT, it is very important to avoid interruptions, since there is a relationship between the delay of the total treatment time (TTT), general survival and the local control rate, resulting in an average loss of the latter from 1.2% per day up to 12-14% per week, requiring an increase in the daily dose of approximately 0.6-0.8 Gy / d to compensate for the extra time. The loss of one or more consecutive sessions in the week has prognostic significance. Only well and moderately differentiated tumors appear to be affected by delays in RT, although poorly differentiated tumors are not affected and depend mainly on the primary tumor^18^.

Nutritional counseling is very important because it is a support in the different phases of treatment to reduce physical deterioration, weight loss and muscle mass. During treatment tissue injuries are generated causing a negative affect in nutrition, for example, chemotherapy damages the epithelium of the digestive tract, about 40% of patients’ present inflammation of the mucosa of the digestive tract, which alters the composition of the intestinal flora. Some drugs can alter taste and smell, such as cisplatin and doxorubicin that cause changes in taste, which interferes with nutrition. Malnutrition in patients with esophageal cancer has been shown to persist for 4 to 6 weeks after cancer treatments, so early detection of the nutritional status of these patients, early referral to dietitians, and effective nutritional interventions are necessary^19^, ^20^.

The Subjective Patient Generated Global Assessment (PG-SGA) is a useful nutritional screening tool, recommended by the American Society for Parenteral and Enteral Nutrition (ASPEN). Nutritional intervention is required when the score is greater than 4 and emergency nutritional intervention is needed when the score is greater than or equal to 9. For patients with esophageal cancer undergoing chemoradiotherapy, with severe or progressive dysphagia before or during treatment, it can indicate the placement of NGT, to avoid interruptions and delays in treatment and it is used for nutritional support <30 days; this procedure is non-invasive, convenient, inexpensive, and can be used when needed. Risks of NGT are esophagitis, reflux, nausea, vomiting, obstruction, ulcer, bleeding, and aspiration pneumonia. For nutritional support> 30 days, PEG is recommended for malnourished patients suffering from inflammation of the oral and esophageal mucosa due to radiotherapy. The PEG is comfortable, has a large diameter which reduces obstructions; risks include; wound infection, mild bleeding, necrotizing fasciitis, and peritonitis^21^.

A retrospective study of patients with squamous cell carcinoma of the esophagus, with malignant fistula, demonstrated that when enteral nutritional support is given with chemoradiotherapy, the fistula can be cured. Furthermore, an overall survival of 25.2 months was estimated in these patients, with 62.5% of patients remaining alive 1 year after treatment^22^. This result indicates that the fistula is not a contraindication for chemoradiotherapy.

In the analysis carried out in this study, it was observed that the group that presented more intra-treatment interruptions were the patients fed EN, with a risk more than 2 times of interrupting than the group that was fed orally, significantly (p <0.05); with more days compared to those who were fed orally, but with a non-significant difference (p> 0.05). The effect of the interruptions was mainly related to the type of nutrition, independent of the other variables. Interruptions occurred more in men, in advanced stages, in which only stage could be a confounding variable.

Dehydration was the most frequent cause of interruption and could be related to anxiety, depression, early satiety, loss of appetite or lack of nutritional advice^2^; secondly, anemia, which may also be due to lack of nutritional monitoring, leading to low protein intake, ignorance of iron absorption facilitators and inhibitors, lack of supplementation; low socioeconomic status or haematological adverse effects of CT and RT. Prospective and retrospective observational trials in patients with inadequate food intake have shown that EN compared to oral feeding reduces weight loss, as well as the frequency and duration of treatment interruptions and rehospitalizations^14^.

According to the results of the present study, the placement of EN prophylactically is not recommended, but if required according to the clinical and nutritional evolution of the patient, to avoid interruptions, following the guidelines of the Chinese consensus of experts in enteral nutrition in esophageal cancer patients treated with radiotherapy^3^. In addition, it has been shown that the insertion of the NGT can cause pain, which can worsen odynophagia^10^.

As limitations of this study, the medical records did not report whether the patients had undergone a nutritional screening such as the PG-SGA in clinical practice or the NRS-2002 for nutritional screening at hospital admission, diagnosis or during treatment. Furthermore, the weight loss of all the patients was not recorded, which is extremely important to establish the most convenient type of nutrition for them^2^, ^21^. The albumin values that allow knowing if the patient is malnourished when the value is <30 g / L were not recorded either; hypoalbuminemia can aggravate metabolic disorders, increase infectious complications, and prolong hospital stay, which is directly related to the poor prognosis of patients and is an independent risk factor that affects prognosis^4,22^.

Much of the limitation of the study is due to the fact that the patients who were evaluated are referred by clinical oncologists from other primary centers and not all of them have an oncological nutrition service, as do the radiotherapy centers; therefore, patients are referred to external nutritionists and consequently there is no standardized multidisciplinary follow-up in the same place. In this context, it is advisable to standardize the use of the aforementioned validated nutritional screening and use them before and during radiation treatment, since nutritional referral is often neglected.

Finally, according to the present study, it is inferred that EN before treatment does not ensure compliance with chemoradiotherapy without interruptions. Future studies are recommended in relation to interruptions during chemoradiotherapy according to the compromised esophageal level and stage, with a larger population to detect whether the interruptions are caused by the type of nutrition or by the disease itself and at the time of treatment. Anamnesis of the patient is important, to inquire more about the previous nutritional follow-up. It also opens the possibility of carrying out future prospective or experimental cohort studies where nutritional screening is established in radiotherapy centers to assess whether it has relevance in the interruptions or if the interruptions are associated with other factors inherent to the disease itself or to the toxicities generated by the treatment.

## Conclusions

Patients with esophageal cancer have a high risk of malnutrition and cachexia, which is exacerbated by the side effects caused by chemoradiotherapy, therefore it is important to perform a nutritional screening in all patients before and during treatment, to avoid interruptions and conditions of the general state of the patient that decrease their quality of life. The delay in the total time of the treatment affects the general survival and the local control rate of the tumors. In this study, it is concluded that the group of patients who underwent EN treatment presented more interruptions during chemoradiotherapy than the patients who were fed orally with a clinically significant difference, so it is recommended that EN not be used prophylactically, only when necessary. It is recommended to carry out this study with a larger population, considering the nutritional data and evolution of the patients. Finally, multidisciplinary work is recommended, including the intervention of nutritionists in favor of the patient’s quality of life during treatment.

## Data Availability

All data produced in the present study are available upon reasonable request to the authors.

